# Analytical and Clinical Validation for RT-qPCR detection of SARS-CoV-2 without RNA extraction

**DOI:** 10.1101/2020.06.24.20134783

**Authors:** José P. Miranda, Javiera Osorio, Mauricio Videla, Gladys Angel, Rossana Camponovo, Marcela Henríquez-Henríquez

## Abstract

**Background:** The recent COVID-19 pandemic has posed an unprecedented challenge to laboratory diagnosis, based on the amplification of SARS-CoV-2 RNA. With global contagion figures exceeding 4 million persons, the shortage of reagents for RNA extraction represents a bottleneck for testing globally. We present the validation results for a RT-qPCR protocol without prior RNA extraction. Because of its simplicity, this protocol is suitable for widespread application in resource-limited settings.

**Methods:** Optimal protocol was selected by comparing RT-qPCR performance under a set of thermal (65°, 70°, and 95° for 5, 10, and 30 minutes) and amplification conditions (3 or 3,5 uL loading volume; 2 commercial RT-qPCR kits with limit of detection below 10 copies/sample) in nasopharyngeal swabs stored at 4°C in sterile Weise’s buffer pH 7.2. The selected protocol was evaluated for classification concordance with the standard protocol (automated RNA extraction) in 130 routine samples and in 50 historical samples with Cq values near to the clinical decision limit.

**Results:** Optimal selected conditions were: Thermal shock at 70° C for 10 minutes, loading 3.5 ul in the RT-qPCR. Prospective evaluation in 130 routine samples showed 100% classification concordance with the standard protocol. The evaluation in historical samples, selected because their Cqs were at the clinical decision limit, showed 94% concordance with our confirmatory-gold standard which includes manual RNA extraction.

**Conclusions:** These results validate the use of this direct RT-qPCR protocol as a safe alternative for SARS CoV-2 diagnosis in case of a shortage of reagents for RNA extraction, with minimal clinical impact.

## INTRODUCTION

In late 2002, an epidemic outbreak of severe acute respiratory syndrome (SARS) was described in China’s Guangdong province and its cause was attributed months later to the SARS-CoV coronavirus (Drosten et al., 2003; Ksiazek et al., 2003; Peiris et al., 2003). According to statistics from the World Health Organization (WHO), this outbreak reached 26 countries, with an estimated number of cases in 8096, of which 774 died (WHO, 2015).

A new strain of coronavirus (SARS-CoV-2), causing COVID-19 disease, was reported in December 2019 in Wuhan, Hubei province, China (Zhu et al., 2020). Since then, this outbreak has spread globally, forcing the WHO to decree a pandemic on March 11^th^, 2020, when the number of confirmed cases reached 118,000 within 114 countries (WHO, 2020b). After 2 months of this decree, many countries find themselves with strict quarantine policies and a number of confirmed cases that globally rise 5.7 million, while death people with COVID-19 reach 357,736 (WHO; May 29, 2020; https://covid19.who.int/).

The rapid availability of the complete genome of this new virus, submitted on January 5^th^ to GeneBank under the access code MN908947 (Wu et al., 2020) and released by January 12^th^, allowed the development of specific primers to amplify the genetic material of SARS-CoV-2 in order to diagnose patients with COVID-19, using the Reverse Transcription Quantitative Real-Time Polymerase Chain Reaction (RT-qPCR) technique. Until May 9^th^, 2020, the Foundation for Innovation in New Diagnostics (FIND; http://www.finddx.org) listed on its website at least 141 different commercially available diagnostic kits for nucleic acid amplification test (NAAT) of SARS-CoV-2 with CE-IVD certification, and this number increases to 314 kits when considering other types of certification for clinical diagnosis. NAAT-based kits, the common analysis for early detection of SARS-CoV-2 (WHO, 2020a), share characteristics in their processing, including 1) Sample collection, typically performed with nasopharyngeal or oropharyngeal swabs 2) RNA extraction and 3) Reverse transcription of RNA, PCR amplification and detection.

Due to the tremendous number of tests that are being carried out globally, reagents necessary for the SARS-CoV-2 detection process are scarce, especially those required for RNA extraction, which represent a dangerous bottleneck to ensure rapid diagnostic process in patients with COVID-19 and its appropriate clinical management (Babiker et al., 2020; Esbin et al., 2020). The aim of this work was to develop and validate a protocol for the detection of SARS-CoV-2 based on RT-qPCR without RNA extraction. The widespread validation and use of this kind of protocol might contribute to ensure diagnostic continuity in the nowadays setting of globally limited resources for manual and automatic viral RNA extraction, helping to outbreak control and characterization.

## MATERIALS AND METHODS

### Clinical specimens

Clinical samples for the standardization and validation experiments were obtained from the routine of ELSA Clinical Laboratories, IntegraMedica, part of Bupa, Santiago, Chile. This laboratory serves annually to 1million patients, with more than 12 million total tests, and corresponds to the biggest private laboratory provider in Chile. Sampling was performed using nasopharyngeal swabs in symptomatic patients and then stored at 4° C in tubes containing sterile potassium sodium phosphate buffer (Weise’s buffer) pH 7.2 (Merck, Cat. No.109468), until analysis.

All procedures followed were in accordance with the Helsinki Declaration. Prior to sampling, all patients requesting COVID-19 PCR testing were asked to approve and sign consent forms allowing the use of their anonymized samples and clinical information for epidemiological vigilance and research.

### Standard RT-qPCR protocol

For the standard protocol, routinely used in the laboratory for the detection of SARS-CoV-2, an aliquot of 180 ul of sample from the primary sample (nasopharyngeal swab or nasopharyngeal aspirates) including 10 ul of extraction control was used to extract RNA with the MagNA Pure 96 DNA and Viral NA LV Kit (Roche Diagnostics, Cat. No. 06374891001) in the MagNA Pure 96 System (Roche Diagnostics). Then, 10 ul of the extracted RNA was used for amplification by RT-qPCR using the LightMix^®^ Modular Wuhan CoV RdRP-gene kit (Roche, Cat. No. 53-0777-96) in a Cobas z 480 system (Roche Diagnostics). This protocol allows the amplification of a 100 bp fragment from a conserved region of the RNA-dependent RNA polymerase (RdRP) gene and was used as a reference for standardization, validation, and final evaluation in problematic samples. Positive, negative, and blank controls (no template or no RT enzyme) were included in all the amplification procedures. Automatic analysis was performed using LightCycler^®^ 480 Software, Version 1.5.

### Direct RT-qPCR Protocol Standardization

For the standardization of the direct SARS-CoV-2 detection protocol without RNA extraction steps, 50 ul aliquots from the primary sample (nasopharyngeal swabs or nasopharyngeal aspirates) of 5 anonymized patients were subjected to heat shock (65°, 70° or 95° C) during different incubation times (5, 10 or 30 minutes), and then were quickly placed at 4° C until the moment of amplification. From the sample subjected to heat shock, 2 different loading volumes were used for the RT-qPCR (3 or 3.5 ul of the sample). The later sample-treatment was evaluated using the LightMix^®^ Modular Wuhan CoV RdRP-gene and the SARS-CoV-2/SARS-CoV Multiplex REAL-TIME PCR Detection Kit (DNA-Technology, Cat. No. R3-P436-23/9EU R3-P436-S3/9EU), using the respective controls, following the manufacturer’s instructions, and loaded in a Roche Cobas z 480 or in a DTlite thermal cycler (DNA-Technology), respectively. Multiplex of DNA-Technology kit amplifies 3 targets, the first is general to SARS-CoV-like coronaviruses (CoV-like), the other 2 targets are specific to SARS-CoV-2, for E gene (CoV-2 E) and N gene (CoV-2 N). Automatic analysis was performed using the DTmaster software for DNA-Technology kit, included in the DTlite system. Quantification cycle (Cq) values obtained with each kit and condition were compared with those obtained with the standard protocol for the same samples.

### Validation of the Direct RT-qPCR Protocol in routine clinical samples

For the validation stage, results obtained for the described sample-treatment conditions and amplification conditions were evaluated by an expert board in the laboratory to select the set of conditions (direct RT-qPCR protocol) with the best performance, considering Cq-value for positive samples and clinical/analytical classification concordance. The following Direct RT-qPCR protocol was selected for further validation: heat shock at 70° C for 10 minutes and then quickly placed at 4° C, loading 3.5 ul of the sample for RT-qPCR amplification with the SARS-CoV-2/SARS-CoV Multiplex REAL-TIME PCR Detection Kit.

This protocol was further evaluated for a) repeatability and analytical variability of Cqs, using an abbreviated protocol that included 4 anonymized clinical samples run in triplicates, b) Statistical and clinical equivalence of the obtained Cqs for positive samples in comparison with the standard protocol already in use (n=27) and c) Clinical classification concordance with the standard protocol already in use (n=130).

### Evaluation of the validated protocol using problematic samples

To test the performance of the direct RT-qPCR protocol in samples with Cqs near or bellow to the discriminatory value (Cq ≥40) as by our standard protocol (denominated “problematic samples” in this text for simplicity), we analyzed 50 historical samples with this condition in parallel by the direct and standard RT-qPCR protocols. In our laboratory routine, we set a confirmation algorithm for these samples, where RNA is manually extracted using RNeasy Mini Kit (Qiagen, Cat. No. 74106) and RT-qPCR amplification is re-run for samples with 1 ng/ul of RNA or more after manual RNA extraction, to establish a final classification. For samples with less than 1 ng/ul RNA after Manual RNA extraction, patients are contacted to take a new sample. Results for the comparison between the Direct RT-qPCR protocol, the standard laboratory protocol (with automatic RNA extraction), and the confirmatory protocol (with manual RNA extraction) are presented.

### Statistics

Descriptive statistics, statistical analyzes, and Bland-Altman graphs for the comparison of the different protocols were performed using Stata MP 14.2.

## RESULTS

Results from the standardization experiment are shown in Table 1. Based on these results and expert laboratory board (MHH, MV, GA, JO), we established the following direct RT-qPCR protocol as optimal for further validation:

**Table 1.** Result of the standardization of optimal conditions for direct RT-qPCR. Different treatment conditions were evaluated with 2 commercial kits and compared with the standard protocol. Amplification on negative or blank samples was not detected.

|  |  |  |  |  |  |  |  |  |  |  |  |  |  |  |
| --- | --- | --- | --- | --- | --- | --- | --- | --- | --- | --- | --- | --- | --- | --- |
| Temperature (°C) |  | 65° | 65° | 70° | **70°** | 95° | 95° |  | 65° | 65° | 70° | **70°** | 95° | 95° |
| Incubation time (min) | - | 30 | 30 | 10 | **10** | 5 | 5 |  | 30 | 30 | 10 | **10** | 5 | 5 |
| Sample volume (ul) |  | 3 | 3.5 | 3 | **3.5** | 3 | 3.5 |  | 3 | 3.5 | 3 | **3.5** | 3 | 3.5 |
|  | Standard Protocol | LightMix® Modular Wuhan CoV RdRP-gene |  |  |  |  |  | SARS-CoV-2/SARS-CoV Multiplex RT-qPCR |  |  |  |  |  |  |
| Anonymized sample | Cq | Cq |  |  |  |  |  | Target | Cq |  |  |  |  |  |
| 4794 | 35.8 | 38.6 | 40.0 | 38.1 | **39.0** | 39.5 | 38.7 | CoV-like | 36.6 | 36.4 | 36.5 | **36.3** | 37.6 | 37.1 |
|  |  |  |  |  |  |  |  | CoV-2 E | 36.7 | 36.5 | 36.6 | **36.3** | 37.7 | 37.2 |
|  |  |  |  |  |  |  |  | CoV-2 N | 36.7 | 37.1 | 36.7 | **36.7** | 38.5 | 37.7 |
| 4793 | 16.2 | 24.5 | 24.5 | 24.1 | **23.2** | 21.6 | 21.8 | CoV-like | 17.4 | 17.2 | 17.4 | **17.3** | 22.6 | 23.6 |
|  |  |  |  |  |  |  |  | CoV-2 E | 17.5 | 17.2 | 17.5 | **17.4** | 22.6 | 23.7 |
|  |  |  |  |  |  |  |  | CoV-2 N | 17.7 | 17.6 | 18 | **17.8** | 22.8 | 23.7 |
| 3023 | (-) | (-) | (-) | (-) | **(-)** | (-) | (-) | CoV-like | (-) | (-) | (-) | **(-)** | (-) | (-) |
|  |  |  |  |  |  |  |  | CoV-2 E | (-) | (-) | (-) | **(-)** | (-) | (-) |
|  |  |  |  |  |  |  |  | CoV-2 N | (-) | (-) | (-) | **(-)** | (-) | (-) |
| 1929 | >40 | (-) | (-) | (-) | **(-)** | (-) | (-) | CoV-like | (-) | (-) | (-) | **(-)** | (-) | (-) |
|  |  |  |  |  |  |  |  | CoV-2 E | (-) | (-) | (-) | **(-)** | (-) | (-) |
|  |  |  |  |  |  |  |  | CoV-2 N | (-) | (-) | (-) | **(-)** | (-) | (-) |
| 2980 | 30.1 | 33.9 | 33.5 | 33.3 | **33.7** | 32.9 | 32.7 | CoV-like | 35.7 | 31.6 | 31.7 | **31.6** | 33.5 | 33.3 |
|  |  |  |  |  |  |  |  | CoV-2 E | 35.8 | 31.8 | 31.7 | **31.7** | 33.7 | 33.3 |
|  |  |  |  |  |  |  |  | CoV-2 N | 36.0 | 31.7 | 31.7 | **31.7** | 33.7 | 33.1 |

1. Obtain an aliquot of 50 ul from the primary sample, stored at 4°C in Weise’s buffer
2. Thermal shock at 70° C for 10 minutes
3. Store at 4° C until loading the sample into a plate
4. Perform the RT-qPCR using 3.5 ul of sample with the SARS-CoV-2/SARS-CoV Multiplex REAL-TIME PCR Detection Kit (high sensibility kit).

A summary of this procedure and a comparison with the standard protocol is detailed in Figure 1.

**Figure 1.**
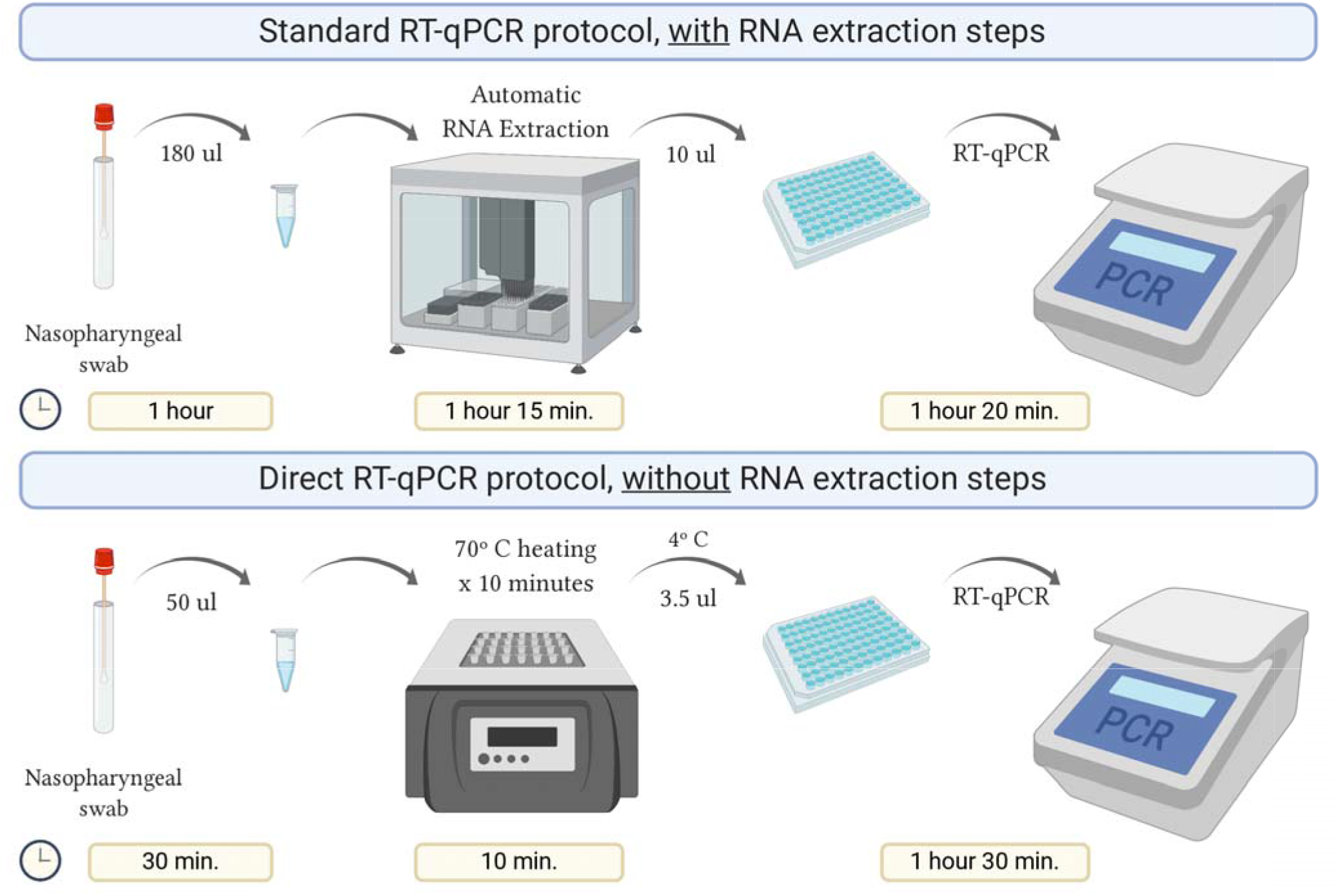
Schematic representation of the direct RT-qPCR protocol without RNA extraction steps, compared to the standard protocol.

Using the direct protocol saved about 40% of the analysis time compared to the standard, which allows enhancing daily processing capability.

Table 2 resumes the results of a brief repeatability assay to roughly characterize the variability of the Cqs. Note that Cqs present a very low intra-assay variability, therefore we proceeded to the validation stage.

**Table 2.** Repeatability and pre-validation of selected RT-qPCR protocol conditions in 4 clinical samples. Incubation time 10 minutes, temperature 70° C, sample volume 3.5 ul using DNA-Technology kit.

|  | Standard Protocol | SARS-CoV-2/SARS-CoV Multiplex RT-qPCR |  |  |  |  |
| --- | --- | --- | --- | --- | --- | --- |
| Anonymized sample | Cq | Target | Cq1 | Cq2 | Cq3 | Informed diagnostic |
| 6608 | (-) | CoV-like<br>CoV-2 E<br>CoV-2 N | (-)<br>(-)<br>(-) | (-)<br>(-)<br>(-) | (-)<br>(-)<br>(-) | Negative |
| 3407 | 26.5 | CoV-like<br>CoV-2 E<br>CoV-2 N | 28.1<br>28.1<br>27.6 | 27.4<br>27.3<br>27.1 | 27.4<br>27.4<br>27.2 | Positive |
| 3380 | 34.3 | CoV-like<br>CoV-2 E<br>CoV-2 N | 36.1<br>36.1<br>36.5 | 35.2<br>35.3<br>35.0 | 37.4<br>37.3<br>37.6 | Positive |
| 3420 | 37.6 | CoV-like<br>CoV-2 E<br>CoV-2 N | 34.3<br>34.4<br>34.3 | 35.2<br>35.2<br>35.5 | 34.5<br>34.6<br>34.9 | Positive |

Diagnostic classification concordance was then evaluated in 130 routine samples, run prospectively using the direct RT-qPCR protocol and the standard protocol. We found full concordance with 27 positive patients for SARS-CoV-2 (Table 3). Moreover, the same was found for the other 103 patients, with negative confirmation by both methods. Based on these results, we established that the performance of the direct protocol was very high, with neither false positive nor false negative results in the 130 samples analyzed, thus yielding to 100% concordance for the 130 samples (Table 5). Additionally, we found that Cqs for positive samples were significantly lower when using the direct protocol and DNA-Technology kit. The median Cq for the standard protocol was 34.1, while for the direct protocol it was 30.4 (CoV-like), 30.6 (CoV-2 E gene), and 30.7 (CoV-2 N gene) (P <0.0009 for each, Wilcoxon signed-rank test) (Table 3). In order to compare this difference, a Bland-Altman graph was made using the standard protocol as a reference (RdRP gene) (Figure 2).

**Table 3.** Validation of the direct RT-qPCR. We evaluated 130 clinical cases, of which 27 were positive (20.8% positivity). Negative samples were omitted from this table.

|  | Standard Protocol | SARS-CoV-2/SARS-CoV Multiplex RT-qPCR |  |  | Informed diagnostic |  |
| --- | --- | --- | --- | --- | --- | --- |
| Anonymized positive samples | Cq RdRP | Cq CoV-like | Cq CoV-2 E | Cq CoV-2 N | Standard Protocol | Direct RT-qPCR |
| 3775 | 28.4 | 29.4 | 29.5 | 30.7 | Positive | Positive |
| 3787 | 25.0 | 20.0 | 21.1 | 20.7 | Positive | Positive |
| 3793 | 32.6 | 29.2 | 29.3 | 29.4 | Positive | Positive |
| 3795 | 39.0 | 30.5 | 30.6 | 31.1 | Positive | Positive |
| 3798 | 29.1 | 28.8 | 28.8 | 28.7 | Positive | Positive |
| 3809 | 26.5 | 23.3 | 23.4 | 23.8 | Positive | Positive |
| 3810 | 21.0 | 22.1 | 22.2 | 22.0 | Positive | Positive |
| 3811 | 37.6 | 32.1 | 32.1 | 32.6 | Positive | Positive |
| 3814 | 34.1 | 32.6 | 32.7 | 33.1 | Positive | Positive |
| 3820 | 27.2 | 24.8 | 24.9 | 24.7 | Positive | Positive |
| 3823 | 35.3 | 30.6 | 30.7 | 31.2 | Positive | Positive |
| 3824 | 36.0 | 30.7 | 30.8 | 30.7 | Positive | Positive |
| 3826 | 35.4 | 32.2 | 32.2 | 32.6 | Positive | Positive |
| 3841 | 32.0 | 30.4 | 30.6 | 30.8 | Positive | Positive |
| 3843 | 25.6 | 22.1 | 22.3 | 22.6 | Positive | Positive |
| 3844 | 25.6 | 28.2 | 28.2 | 27.7 | Positive | Positive |
| 3846 | 35.2 | 36.5 | 36.6 | 38.2 | Positive | Positive |
| 3852 | 28.9 | 29.3 | 29.3 | 29.4 | Positive | Positive |
| 3854 | 31.5 | 26.2 | 26.3 | 26.4 | Positive | Positive |
| 3855 | 35.4 | 27.6 | 27.6 | 28.0 | Positive | Positive |
| 3859 | 33.2 | 28.8 | 29.0 | 29.7 | Positive | Positive |
| 3865 | 37.0 | 33.9 | 33.9 | 34.3 | Positive | Positive |
| 4890 | 39.0 | 39.0 | 39.1 | 40.4 | Positive | Positive |
| 5003 | 39.2 | 34.3 | 34.4 | 34.7 | Positive | Positive |
| 5174 | 38.7 | 35.7 | 35.8 | 36.6 | Positive | Positive |
| 9107 | 39.3 | 37.1 | 37.0 | 36.6 | Positive | Positive |
| 13848 | 39.3 | 39.2 | 39.2 | 39.8 | Positive | Positive |
| **Median Cq (IQR)** | 34.1 (28.4-37.6) | 30.4 (27.6-33.9) | 30.6 (27.6-33.9) | 30.7 (27.7-34.3) |  |  |
| **P-value** | - | **0.0002\*** | **0.0003\*** | **0.0009\*** |  |  |

\*Wilcoxon signed-rank test, Cq values from standard protocol used as reference. IQR = inter quartile range.

**Table 5.** Evaluation of clinical performance of direct RT-qPCR protocol compared to standard protocol as reference. Concordance for each, the validation and evaluation of problematic samples are shown.

|  | Validation Stage (Cq <40) | Evaluation of Problematic Samples (Cq ≥40) |
| --- | --- | --- |
| True Positive | 27 | 31 |
| True Negative | 103 | 16 |
| False Positive | 0 | 2 |
| False Negative | 0 | 1 |
| Total | 130 | 50 |
| Classification concordance | 100% | 94% |

**Figure 2.**
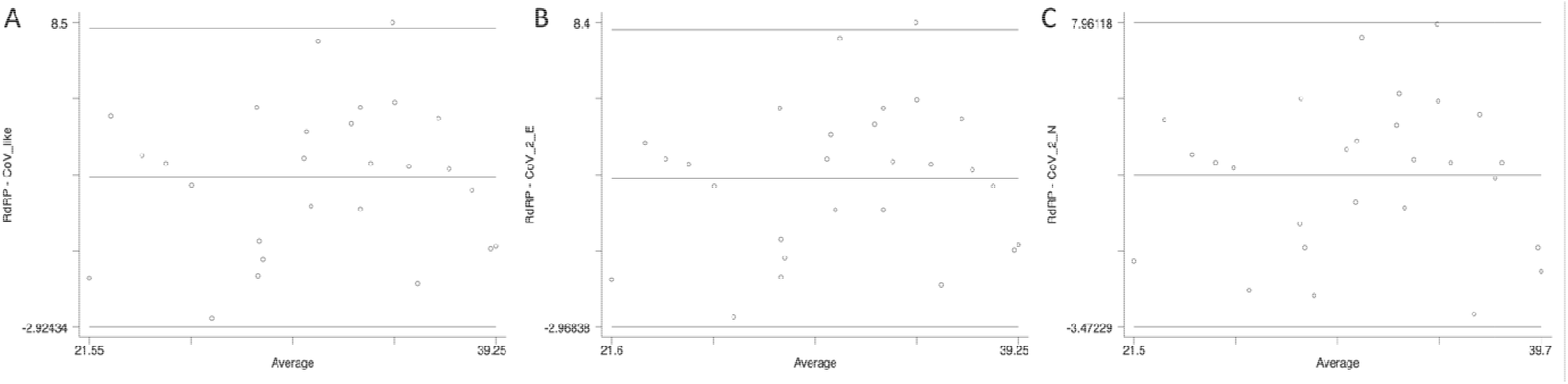
Bland-Altman comparisons between Cq values obtained for RdRP gene under standard protocol and A) CoV-like, B) Cov-2 E-gene and C) CoV-2 N-gene using DNA-Technology kit.

With the purpose of evaluating the performance of direct RT-qPCR protocol in samples with Cq close to the discriminatory value, a total of 50 historical samples analyzed with the standard protocol and confirmed by manual extraction according to our current quality assurance algorithm were re-analyzed by direct RT-qPCR (Table 4). As observed, there is a high classification agreement with the confirmatory protocol that reaches 94% (Table 5), which is also observed in terms of Cq (Table 4). The only discordance is 2 samples classified as positive with the direct RT-qPCR protocol (possibly false positive), while they showed amplification over cycle 40 when they were processed by automatic and manual RNA extraction. We also obtained a sample with amplification after cycle 40 with the direct method, previously classified as positive according to the standard method (possibly false negative). In every case, direct RT-qPCR presents amplification close to cycle 40, so it is plausible that the sample corresponded to patients with a viral load very close to the detection limit for both protocols.

**Table 4.** Analysis of concordance in classification of 50 problematic historical samples, which were re-processed using the direct RT-PCR validated protocol. For standard protocol, classification was performed using results from manual RNA extraction.

|  | Standard Protocol | Standard Protocol (manual extraction) | Direct SARS-CoV-2/SARS-CoV Multiplex RT-qPCR |  |  | Informed diagnostic |  |
| --- | --- | --- | --- | --- | --- | --- | --- |
| Anonymized samples | Cq RdRP | Cq RdRP | Cq CoV-like | Cq CoV-2 E | Cq CoV-2 N | Standard Protocol (manual) | Direct RT-qPCR |
| 604 | >40 | >40 | (-) | 40.3 | 39.2 | Negative | Negative |
| 189 | >40 | 34.3 | 38.3 | 38.6 | 38.4 | Positive | Positive |
| 193 | >40 | 33.0 | 36.0 | 36.0 | 36.3 | Positive | Positive |
| 605 | >40 | 34.1 | 32.4 | 32.4 | 32.9 | Positive | Positive |
| 624 | >40 | >40 | 40.4 | 40.3 | 39.2 | Negative | Negative |
| 650 | >40 | >40 | 36.1 | 36.1 | 36.9 | **Negative** | **Positive** |
| 727 | >40 | >40 | 39.7 | 38.9 | 39.8 | Negative | Negative |
| 1265 | >40 | 36.3 | 35.9 | 35.9 | 36.7 | Positive | Positive |
| 1287 | >40 | 28.6 | 26.7 | 26.8 | 27.2 | Positive | Positive |
| 1288 | >40 | >40 | (-) | (-) | (-) | Negative | Negative |
| 1298 | >40 | 26.5 | 25.7 | 25.5 | 25.0 | Positive | Positive |
| 1309 | >40 | 29.9 | 28.2 | 28.1 | 28.7 | Positive | Positive |
| 1346 | >40 | 28.5 | 28.2 | 28.3 | 28.3 | Positive | Positive |
| 1421 | >40 | (-) | 38.9 | 38.9 | (-) | Negative | Negative |
| 1433 | >40 | 30.0 | 28.9 | 28.8 | 30.1 | Positive | Positive |
| 1434 | >40 | 37.0 | 35.3 | 35.3 | 36.7 | Positive | Positive |
| 1576 | >40 | 31.3 | 29.2 | 29.1 | 29.4 | Positive | Positive |
| 1637 | >40 | (-) | (-) | (-) | (-) | Negative | Negative |
| 1684 | >40 | 30.8 | 29.5 | 29.5 | 30.4 | Positive | Positive |
| 1708 | >40 | 34.9 | 33.3 | 33.3 | 34.5 | Positive | Positive |
| 1726 | >40 | (-) | (-) | (-) | (-) | Negative | Negative |
| 1734 | >40 | 36.1 | 33.9 | 33.9 | 35.0 | Positive | Positive |
| 1742 | >40 | 33.9 | 32.4 | 32.4 | 32.3 | Positive | Positive |
| 1792 | >40 | (-) | (-) | (-) | (-) | Negative | Negative |
| 1802 | >40 | (-) | (-) | (-) | (-) | Negative | Negative |
| 1850 | >40 | 24.9 | 24.2 | 24.4 | 25.0 | Positive | Positive |
| 1883 | >40 | 34.0 | 32.6 | 32.6 | 32.7 | Positive | Positive |
| 1921 | >40 | 32.9 | 32.3 | 32.3 | 31.8 | Positive | Positive |
| 1929 | >40 | >40 | 37.9 | 37.8 | 38.2 | **Negative** | **Positive** |
| 1968 | >40 | 30.2 | 28.4 | 28.4 | 29.6 | Positive | Positive |
| 1983 | >40 | 30.6 | 29.1 | 29.2 | 29.7 | Positive | Positive |
| 1997 | >40 | (-) | (-) | 40.9 | (-) | Negative | Negative |
| 3440 | >40 | 29.3 | 33.7 | 33.8 | 33.4 | Positive | Positive |
| 3445 | >40 | 35.5 | 36.1 | 36.2 | 36.7 | Positive | Positive |
| 3479 | >40 | 31.1 | 31.7 | 31.8 | 32.0 | Positive | Positive |
| 3484 | >40 | (-) | (-) | (-) | (-) | Negative | Negative |
| 3507 | >40 | (-) | (-) | (-) | (-) | Negative | Negative |
| 3539 | >40 | 34.7 | 35.9 | 36.0 | 35.9 | Positive | Positive |
| 3587 | >40 | >40 | (-) | (-) | 41.4 | Negative | Negative |
| 3602 | >40 | 35.6 | 39.9 | 39.8 | 38.6 | Positive | Positive |
| 3654 | >40 | (-) | (-) | (-) | (-) | Negative | Negative |
| 4167 | >40 | 35.9 | 37.9 | 37.9 | 38.6 | Positive | Positive |
| 4168 | >40 | 29.8 | 37.4 | 37.5 | 37.8 | Positive | Positive |
| 4169 | >40 | 34.4 | 35.5 | 35.6 | 36.0 | Positive | Positive |
| 4170 | >40 | 33.5 | 36.5 | 36.5 | 36.5 | Positive | Positive |
| 4174 | >40 | (-) | (-) | (-) | (-) | Negative | Negative |
| 4183 | >40 | >40 | (-) | (-) | (-) | Negative | Negative |
| 7226 | >40 | 38.0 | 41.9 | 42.0 | 46.4 | **Positive** | **Negative** |
| 7626 | >40 | 30.5 | 33.6 | 33.7 | 33.8 | Positive | Positive |
| 8259 | >40 | 37.3 | 37.7 | 38.5 | 38.1 | Positive | Positive |

## DISCUSSION

The evidence presented allows validating the direct RT-qPCR protocol as a comparable alternative to the standard protocol in routine for detection of SARS-CoV-2. Consequently, the clinical impact of replacing the standard protocol currently in use in our laboratory with the new direct RT-qPCR protocol is estimated to be minimal, given the high classification agreement between both techniques, without false negatives and false positives in a total of 130 samples analyzed by both methods as part of the described validation stage.

In a small number of samples using the standard protocol in our laboratory routine, clear amplification is obtained after cycle 40. We have denominated these samples as “problematic” because it is difficult to establish whether it is a true or a false negative. In our experience with the routine use of the Roche LightMix^®^ Modular Wuhan CoV RdRP-gene kit associated with automatic nucleic acid extraction in the MagNA Pure 96 system, approximately 64% of the abovementioned samples (which should be reported as negative using the standard protocol) changed their classification when repeated by manual extraction (Table 4), evidencing the loss of sensitivity in cases with low viral load when performing nucleic acid extraction on an automatic platform. Consequently, we evaluated the direct protocol in 50 samples in this situation and found 94% of concordance with the confirmatory protocol (manual extraction followed by amplification using LightMix^®^ Modular Wuhan CoV RdRP-gene kit) as the gold standard, with only 2 false positive results and 1 false negative result for the direct RT-qPCR. Even when these results are far superior to results obtained by our standard protocol, we conclude that the direct protocol should be evaluated with caution when there is amplification near to the detection limit and thus we decided to set a confirmatory protocol for samples with Cqs ≥37 by the direct method and to re-analyzed these samples using manual RNA extraction and a different RT-qPCR kit.

The demand for tests for the detection of SARS-CoV-2 by RT-qPCR grows every day around the world to allow the management of COVID-19 disease. Despite the high number of commercially available NAA tests, the process requires various reagents and supplies that have become the bottleneck for rapid analysis. Among these reagents, undoubtedly the most scarce have been those related to the extraction of genetic material from the coronavirus. In order to cope with this problem, many research laboratories have joined in the duty of analyzing clinical samples from patients in several countries, playing a fundamental role in maintaining the diagnostic process and containing the spread of the coronavirus, notwithstanding the stocks that are reserved in these centers are also limited and do not allow the long-term diagnostic process. Taking this background into account, we believe that it is vitally important that clinical laboratories take an active role in generating knowledge to maintain the diagnostic process and to share this knowledge, so that others can also implement it.

In the present work, we successfully validated a protocol for performing RT-qPCR bypassing the initial nucleic acid extraction step with a high classification agreement with our standard protocol. So far, a protocol with similar conditions for sample treatment can be found in the literature (Alcoba-Florez et al., 2020), nonetheless, researchers have found that its use required on average 6.1 (± 1.6) more cycles to reach the diagnosis, which is not optimal for the use in clinical laboratory due to the susceptibility of false negatives near to the detection limit. The amplification kit used in their protocol was the LightMix^®^ Modular Wuhan CoV E-gene (Roche Diagnostics), which has a reported analytic sensibility of 10 copies or fewer, performance confirmed by another independent laboratory (Okamaoto et al., 2020). Considering these authors also used a high sensitivity kit, we believe therefore that components of the medium used for swab storage (VTM) could have influenced the displacement of the number of cycles required for the diagnosis. In our protocol, samples were stored in Weise’s buffer, allowing low interference with RT-qPCR. Moreover, our selected amplification kit was manufactured by DNA-Technology and also reports an analytical sensitivity of 10 copies, which agrees with the high classification performance we obtained.

Finally, we consider whether our protocol is extendable to other kits, including those with intermediate sensitivity. With this question in mind, we used the Genesig® kit (Cat No. Z-Path-2019-nCoV, analytical sensitivity <100 copies) on 58 samples. The optimal conditions for direct RT-qPCR protocol in this case were different, with heat shock at 95° C for 5 minutes and loading 6.5 ul of sample. With this protocol, we obtained 23 positive and 35 negative samples, all with a correct diagnostic classification when compared with our Roche standard. The median value for Cq obtained by Roche was 27.2 (IQR 24.3-33.8), while for the Genesig® kit this value was significantly lower, with a median of 25.0 (IQR 24.0-30.8), P = 0.002, Wilcoxon signed-rank test (Data not shown). Considering these data, it is possible to conclude that the most important factor for the success of a direct protocol is to use a sample storage buffer that does not interfere with the RT-qPCR reaction. Furthermore, it strongly suggests that the optimal conditions for use with the direct protocol must be previously tested for each kit.

Despite the fact that this protocol allows to considerably reduce the processing time, we believe that its implementation should be restricted only to those clinical laboratories in which the lack of RNA extraction reagents is a limiting factor to comply with the diagnostic process. This is because the most important object is to ensure the quality of analysis in the diagnosis of patients. Consequently, we hope that the use of this protocol will contribute to ensuring the diagnostic process both in Chile and in other countries.

## Data Availability

All relevant information has been included in the article

## Ethics Statement

All procedures followed were in accordance with the Helsinki Declaration. Clinical samples were anonymized and all patients signed out Informed Consent allowing the use of their samples for research.

### Acknowledgments

We want to express our respect and admiration to all the medical and collaboration personnel who unite day by day to win the battle against the COVID-19 pandemic.

## Author Contributions

All the aforementioned authors have made a substantial contribution to the development of this work. Specifically, JPM wrote the manuscript with the help of MHH. JPM, JO, MV and GA developed the analysis of samples and statistics. RC contributed to the interpretation of the results. MHH conceived the study and was in charge of the direction and planning.

